# The VOICE-DEP study protocol: multimodal analysis of voice and discourse during medical interviews to support diagnosis and longitudinal monitoring of major depressive disorder

**DOI:** 10.64898/2026.07.01.26357063

**Authors:** Maialen Zabalza-Zudaire, Onintza Sayar-Beristain, Paula Fructos, Favio Emanuel Núñez, Franco Fabricio Carpio, Esmeralda García, Álvaro Ortiz, Felipe Ortuño, Azucena Aldaz, Patricio Molero

**Author notes:** Corresponding authors: Maialen Zabalza-Zudaire. Department of Mathematics and Statistics, NNBi 2020 S.L., Galar (Spain)., Patricio Molero, MD, PhD. Department of Psychiatry and Clinical Psychology. Clínica Universidad de Navarra. Av. Pío XII, 36. 31008 Pamplona (Spain). These authors contributed equally to this work (Azucena Aldaz, Patricio Molero).

## Abstract

**Background:** Major depressive disorder is a severe, recurrent and disabling condition. Although diagnosis and clinical monitoring are based on medical interviews and validated rating scales, speech and discourse analysis may provide complementary digital biomarkers reflecting depressive severity and clinical evolution. However, current evidence remains limited by methodological heterogeneity, predominantly cross-sectional designs, limited longitudinal data and underrepresentation of non-English-speaking clinical populations.

**Objective:** The aim of the VOICE-DEP study is to develop and formalize a standardized, reproducible and clinically grounded protocol for the multimodal analysis of voice and discourse during medical interviews as a tool to support the diagnosis of depressive disorder and to assess whether speech-derived biomarkers change over time in parallel with clinical severity measures.

**Methods:** VOICE-DEP is an observational, prospective, longitudinal pilot study of patients with major depressive disorder with a healthy control group, conducted in a hospital-based clinical setting in Spain. The study will include 25 adult patients with moderate or severe unipolar depression, with or without psychotic symptoms, and 50 healthy controls without a personal history of psychiatric disorders. Patients will be assessed at five time points: baseline (V0) and four monthly follow-up visits at 30, 60, 90 and 120 days. Healthy controls will be assessed once at baseline. The planned dataset comprises 175 voice recordings: 125 from patients and 50 from controls. At each assessment, the Montgomery-Asberg Depression Rating Scale related part of the medical interview, lasting approximately 10–30 minutes and including an initial free-speech segment, will be recorded using a standardized audio protocol. Acoustic, paralinguistic and linguistic features will be extracted and analyzed in relation to clinician-rated severity measures and self-reported symptoms.

**Ethics:** This protocol has been reviewed and approved by the local Research Ethics Committee, which complies with the international standards of GCP CPMP/ICH/135/95 (Comunidad Foral de Navarra Research Ethics Committee; reference code: 2026.110). Written informed consent will be obtained from all participants before any study procedure. Voice recordings and clinical data will be pseudonymized, stored securely and processed in accordance with applicable Spanish and European data protection regulations.

**Expected outcomes:** This protocol is expected to generate a clinically grounded Spanish-language longitudinal speech corpus and a transparent analytical framework for evaluating voice- and discourse-derived biomarkers as complementary tools for depression assessment and monitoring.

## 1. Introduction

Major depressive disorder (MDD) is one of the leading contributors to global disability, affecting more than 280 million people worldwide and constituting a major public health challenge [1]. Despite decades of research, clinical assessment and monitoring of depression still rely predominantly on structured interviews and rating scales, which are inherently subjective and susceptible to recall bias and inter-rater variability [2,3].

In recent years, the identification of objective, scalable and non-invasive biomarkers has become a central goal of precision psychiatry [4]. Among behavioral signals, human speech has emerged as a promising source of information because speech production integrates motor control, cognition, affective regulation and autonomic function [5]. Alterations in prosody, speech rate, pauses, intensity and voice quality have been consistently associated with depressive states [6,7].

Recent advances in artificial intelligence have accelerated research on voice-based depression assessment. Machine learning, natural language processing and large language models have shown promising results using acoustic, paralinguistic and linguistic features extracted from speech recordings [8,9]. In parallel, recent studies have increasingly adopted multimodal approaches integrating audio and text information. Representative studies in the field are summarized in Table 1.

**Table 1.** Representative state-of-the-art studies in voice-based depression research.

| Reference | Design and Dataset | Sample (N) |
| --- | --- | --- |
| Jin et al., 2026 [14] | Cross-sectional; MMPsy adolescent dataset (text + audio, PHQ-9). | 1275 participants |
| Li et al., 2025 [15] | Cross-sectional; AVEC2013 and AVEC2014 speech datasets (BDI-II). | 84 subjects (150 clips) |
| Xu et al., 2025 [16] | Cross-sectional; CMDC and DAIC-WOZ multimodal datasets (voice + text, PHQ-8). | 234 subjects |
| Ali et al., 2025 [17] | Longitudinal; DEW dataset (multimodal speech). | Not reported |
| Mazur et al., 2025 [18] | Cross-sectional; remote personal devices. | 14,898 |
| Zhang et al., 2024 [9] | Cross-sectional; DAIC-WOZ dataset (text + acoustic landmarks). | 189 interviews |
| Ghosh et al., 2024 [19] | Cross-sectional controlled; DAIC-WOZ. | 189 participants |
| Huang et al., 2024 [20] | Cross-sectional controlled; DAIC-WOZ. | 189 participants |
| Sadeghi et al., 2024 [21] | Cross-sectional; E-DAIC dataset (text + facial features, PHQ-8). | 275 interviews |
| Anand et al., 2024 [22] | Cross-sectional; E-DAIC dataset (text, audio and visual; PHQ-8). | 275 interviews |
| Menne et al., 2024 [23] | Cross-sectional; clinical cohort using speech tasks (BDI-II / HAMD). | 96 participants |
| Ronneberg et al., 2024 [24] | RCT-controlled; voice-based AI coaching. | 200 participants |
| Wu et al., 2023 [8] | Cross-sectional; DAIC-WOZ dataset using SSL speech representations. | 189 interviews |
| Cansel et al., 2023 [25] | Cross-sectional controlled; smartphone-based recordings. | 104 participants |
| Berardi et al., 2023 [26] | Cross-sectional controlled; digital voice recorder. | 60 participants |
| Li et al., 2023 [27] | Cross-sectional; clinical interviews with NLP-based analysis. | 329 participants |
| Kim et al., 2023 [28] | Cross-sectional controlled; smartphone-based recordings. | 318 participants |
| Wasserzug et al., 2023 [29] | Cross-sectional controlled; mobile phone recordings. | 144 participants |

HC = Healthy Controls; MDD = Major Depressive Disorder; SZ = Schizophrenia; BD = Bipolar Disorder; ANX = Anxiety Disorder; RCT = Randomized Controlled Trial; LLM = Large Language Model; SSL = Self-Supervised Learning; NLP = Natural

However, systematic reviews continue to highlight substantial methodological heterogeneity across this literature [10]. Differences in recording conditions, speech tasks, feature extraction pipelines, validation strategies and clinical reference standards limit reproducibility and comparability across studies. Moreover, the current literature remains dominated by cross-sectional designs and English-speaking datasets, while longitudinal studies in clinically characterized Spanish-speaking populations remain scarce.

Another important limitation is that many studies rely primarily on self-report questionnaires as reference labels for model training and evaluation. In contrast, clinician-rated instruments such as the Montgomery–Asberg Depression Rating Scale (MADRS) may provide a more clinically grounded severity reference given its demonstrated validity and inter-rater reliability [11]. Longitudinal designs are also particularly relevant because they allow evaluation of whether speech-derived biomarkers evolve in parallel with changes in depressive severity over time [12].

Recent evidence in Spanish-speaking clinical populations further supports the relevance of integrating linguistic information into speech-based depression assessment [13]. Nevertheless, important challenges remain unresolved, including the standardization of recording conditions, harmonization of clinical anchors, definition of speech elicitation procedures and longitudinal evaluation of within-subject changes. These limitations highlight the need for transparent and reproducible protocol-driven studies designed specifically for clinical translation.

The present study protocol addresses these challenges by proposing a longitudinal, hospital-based framework for the collection and analysis of speech data in Spanish-speaking patients with depressive disorder and healthy controls. By integrating acoustic, paralinguistic and linguistic features and anchoring them to validated clinician-administered scales, the VOICE-DEP study aims to advance voice-based depression research toward clinically interpretable and reproducible methodology.

## 2. Materials and methods

### 2.1 Aim of the study, design and setting

The overall aim of the VOICE-DEP study is to develop and formalize a standardized, reproducible and clinically grounded protocol for the multimodal analysis of voice and discourse during medical interviews as a tool to support the diagnosis and longitudinal monitoring of depressive disorder.

The working hypothesis is that major depressive disorder and its clinical evolution are associated with changes in acoustic, paralinguistic and linguistic characteristics of voice and discourse, and that these changes can be analyzed in a standardized and reproducible manner in a routine clinical setting.

#### Primary objective

The primary objective is to establish a reproducible clinical framework for extracting and analyzing acoustic, paralinguistic and linguistic features from semi-structured medical interviews, allowing discrimination between patients with a medical diagnosis of depressive disorder and healthy controls using voice-based models anchored to validated clinician-rated clinical reference standards.

#### Secondary objectives

The secondary objectives are: i) To construct a longitudinal Spanish-language clinical speech corpus including repeated voice recordings from patients with depressive disorder and single-session recordings from healthy controls; ii) To characterize the association between speech-derived digital biomarkers and clinician-rated measures of depressive severity, including their evolution over time; iii) To evaluate the added value of multimodal models combining acoustic, paralinguistic and linguistic features compared with unimodal models; iv) To assess intra-individual vocal changes across repeated assessments and their association with changes in clinical severity; v) To provide a transparent methodological framework that may facilitate replication, cross-study comparison and future external validation in other clinical presentations, diagnoses, languages and sociocultural settings; vi) To explore whether specific treatment modalities are associated with greater changes in voice and discourse in parallel with clinical evolution.

#### Study design and setting

This is an observational, prospective, longitudinal pilot study with a healthy control group. The study will be conducted in a hospital-based clinical setting, including both outpatient consultations and inpatient care at the Department of Psychiatry of a university hospital. The planned study duration is 24 months.

### 2.2 Study population and participant characteristics

The study will include two groups: (1) patients with a medical diagnosis of moderate or severe unipolar depression and (2) healthy control participants without a personal history of depression or any other psychiatric disorder.

Inclusion criteria for patients are: Age 18 years or older; Native Spanish speakers or individuals with sufficient fluency in Spanish to maintain a clinical interview; Capacity to understand study procedures and provide written informed consent, according to the investigator’s judgement; Medical diagnosis of moderate or severe unipolar depression, with or without psychotic symptoms, according to ICD-10 categories: moderate depressive episode (F32.1), severe depressive episode without psychotic symptoms (F32.2), severe depressive episode with psychotic symptoms (F32.3), recurrent depressive disorder, current episode moderate (F33.1), recurrent depressive disorder, current episode severe without psychotic symptoms (F33.2), or recurrent depressive disorder, current episode severe with psychotic symptoms (F33.3), regardless of whether the patient receives psychotherapeutic, pharmacological or neurostimulation treatment, or chooses not to receive active treatment while remaining under clinical follow-up.

Exclusion criteria for patients are: Severe neurological disorders affecting speech production or cognitive function, such as severe dementia; Severe voice or speech disorders unrelated to depression that prevent completion of the clinical interview through verbal language; State of intoxication due to substance use at the time of assessment; Other severe medical conditions interfering with study participation or speech production.

Inclusion criteria for healthy controls are: Age 18 years or older. Exclusion criteria for healthy controls are: Personal history of depression or any other psychiatric disorder; Severe neurological disorders affecting speech production or cognitive function, such as severe dementia; Severe voice or speech disorders unrelated to depression that prevent completion of the clinical interview through verbal language; State of intoxication due to substance use at the time of assessment; Other severe medical conditions interfering with study participation or speech production.

### 2.3 Study procedures and visit schedule

Participants will be recruited consecutively according to eligibility criteria. Patients will be invited by physicians from the research team during routine clinical practice. Healthy controls will be invited by members of the research team and, if they agree to participate, will be referred to a physician from the research team for baseline assessment. As this is an observational study, no randomization or blinding procedures will be applied.

At the baseline visit (V0), all participants will receive verbal and written information about the study objectives, methodology, non-invasive nature of the procedures, confidentiality safeguards, and exclusive research use of the collected data. Written informed consent will be obtained before any study-related procedure. A clinical interview will then be conducted to collect baseline sociodemographic and clinical information, followed by psychometric assessment using the clinical instruments specified in the protocol. During the same session, voice recordings will be obtained under standardized conditions.

Only patients with depressive disorder will undergo longitudinal follow-up. Patients will attend four additional visits coinciding, whenever possible, with routine psychiatric follow-up visits: V1 at 30 days, V2 at 60 days, V3 at 90 days and V4 at 120 days, each with an allowed window of ±7 days. At each follow-up visit, the same recording and clinical assessment procedures applied at baseline will be repeated. Clinician-rated and self-reported measures of depressive severity will be updated, and relevant clinical changes, including treatment adjustments or adverse events, will be documented.

Healthy controls will complete only the baseline visit under the same recording conditions. In all participants, data collection will be integrated into the clinical interview. Only the part of the medical interview corresponding to the MADRS assessment will be audio recorded. This recorded segment is expected to last approximately 10–30 minutes and will include an initial free-speech segment, allowing speech and discourse to be collected within a clinically meaningful interaction while maintaining a standardized procedure.

Voice recordings will be assigned a pseudonymized audio identifier linked to participant type, participant code and visit number. Any recording incident, including interruption due to patient need, clinical need or technical problems, will be documented in the case report form and considered in data quality assessment.

### 2.4 Sample size and sampling strategy

The planned study sample comprises 75 participants: 25 patients with a clinical diagnosis of moderate or severe unipolar depression and 50 healthy controls. Patients will be assessed at five time points: baseline (V0) and four monthly follow-up visits at 30, 60, 90 and 120 days, each with an allowed window of ±7 days. Assuming complete follow-up, this will generate 125 longitudinal voice recordings in the clinical group. Healthy controls will undergo a single baseline assessment, generating 50 voice recordings. The planned dataset therefore comprises 175 voice recordings.

The sample size was defined pragmatically as a feasible recruitment target for a hospital-based pilot methodological validation study conducted in a real-world clinical setting. The study is not intended as a definitive diagnostic accuracy trial powered to estimate the performance of a final clinical classifier. Rather, it is designed to evaluate the feasibility, standardization, reproducibility and analytical validity of a multimodal voice- and discourse-based methodology for depression assessment and longitudinal monitoring.

The sampling strategy will be consecutive and non-probabilistic. Eligible patients will be recruited during routine clinical care in the Department of Psychiatry, including both outpatient consultations and inpatient care. Healthy controls will be invited by members of the research team and assessed after confirmation of eligibility.

Although a balanced baseline comparison between patients and healthy controls would be methodologically desirable, the final allocation was defined according to feasibility considerations. Healthy controls will match age and sex of patients recruited as much as possible. Recruitment of clinically characterized patients with moderate or severe depressive disorder is expected to be more demanding because it requires repeated clinical assessments and longitudinal follow-up. In contrast, healthy controls require only a single baseline visit, allowing the inclusion of a larger control group without substantially increasing study burden. Therefore, the study will include 25 patients and 50 healthy controls. This design is expected to strengthen the baseline characterization of non-depressed speech patterns while preserving the longitudinal focus of the clinical cohort.

At participant level, baseline analyses will compare patients and healthy controls, whereas longitudinal analyses will focus on within-patient changes across repeated assessments. For predictive modeling, the imbalance between groups will be explicitly considered through appropriate analytical strategies, such as participant-level data splitting, balanced resampling, class weighting or sensitivity analyses when applicable.

Because repeated recordings from the same patient are not independent, all longitudinal analyses and validation procedures will account for within-subject correlation. For predictive modeling, recordings from the same participant will not be split across training and test sets, in order to reduce optimistic bias and avoid information leakage.

Given the pilot nature of the study and the moderate sample size, classifier performance estimates will be interpreted as exploratory and hypothesis-generating. The study will prioritize transparent feature extraction, reproducible preprocessing, clinically interpretable modeling and longitudinal within-subject analysis over the development of a definitive deployable diagnostic model.

### 2.5. Voice recording protocol

Voice recordings will be obtained using a Jabra Speak 510 USB omnidirectional microphone, positioned frontally at a distance of no more than 20 cm from the participant’s mouth. Recordings will be conducted in a quiet hospital room, with minimized background noise, closed doors and windows whenever feasible, silenced electronic devices, and no overlapping speech or external interruptions whenever possible.

The Jabra Speak 510 microphone was selected for pragmatic and translational reasons: it is affordable, widely available, easy to deploy and compatible with routine hospital use. This choice prioritizes ecological validity and future scalability over laboratory-grade audio fidelity, while maintaining a standardized recording procedure. This approach is consistent with previous studies that have used non-specialized recording devices, including tablet microphones, smartphone microphones and mobile phones, to extract clinically relevant speech and voice features [23,28–30].

Efforts will be made to keep recording times as consistent as possible, as vocal parameters may exhibit circadian fluctuations. The exact recording time will be logged and considered as a prespecified sensitivity factor.

Audio recordings will be stored in uncompressed WAV format to preserve signal integrity during feature extraction. All files will be labeled using pseudonymized alphanumeric identifiers. During preprocessing, interviewer speech will be removed, and only the participant’s speech signal will be retained for analysis.

### 2.6 Variables

All study variables will be recorded in a pseudonymized case report form developed specifically for the VOICE-DEP study. Variables will be classified into: (i) baseline variables collected only at V0, and (ii) longitudinal visit variables collected at V0, V1, V2, V3 and V4 in patients with depressive disorder. Healthy controls will complete only V0.

Exposure variable: The main exposure variable is the medical diagnosis of moderate or severe unipolar depressive disorder, with or without psychotic symptoms, according to the predefined ICD-10 categories included in the protocol.

Primary outcome variables: The primary outcome variables are the standardized voice recording and the multimodal speech- and discourse-derived variables extracted from the recorded clinical interview. These variables will be grouped into acoustic, paralinguistic and linguistic domains.

Clinician-rated and self-reported symptom variables: At each study visit, depressive symptom severity and global clinical status will be assessed concurrently with the voice recording using the MADRS, the Clinical Global Impression scale (ICG), and the Patient Health Questionnaire-9 (PHQ-9). These variables will provide the clinical reference outcomes for cross-sectional and longitudinal analyses.

Baseline and longitudinal variables: Baseline variables collected at V0 will include demographic, anthropometric, clinical and contextual information, including psychiatric and medical history, substance use, stressful life events, and previous depressive episodes. Longitudinal variables collected across visits will include recording traceability variables, symptom scales, pharmacological treatment variables, psychotherapy variables and neurostimulation-related variables.

Metadata related to the recording process, including pseudonymized audio identifiers and recording incidents, will also be documented for traceability and quality control purposes.

The complete list of study variables operationalized in the VOICE-DEP case report form is summarized in Table 2. The list of multimodal speech- and discourse-derived variables is summarized in Table 3.

**Table 2.** Study variables recorded in the VOICE-DEP case report form and assessment schedule.

| Domain | Description | Assessment visits |
| --- | --- | --- |
| Participant identification | Participant category (patient or control), study code, and visit date | V0–V4 in patients; V0 in controls |
| Sociodemographic and anthropometric baseline variables | Sex, age, marital status, place of residence, birth order in the sibship, height, weight, body mass index, body surface area, educational level, childcare responsibilities, caregiving responsibilities for dependent relatives, religious belief, community religious practice, and social relationships | V0 only |
| Lifestyle, family history and contextual baseline variables | First-degree family psychiatric history, toxic habits, tobacco, alcohol, cannabis, cocaine, amphetamine and other substance use, physical activity, diet pattern, complementary or alternative treatments and specific prescribed diets, pharmacogenetic or pharmacodynamic isoform information when available, employment status, night work, acute and chronic stressful life events, and current and lifetime traumatic events | V0 only |
| Clinical baseline variables | Principal diagnosis according to ICD-10 depressive categories, psychiatric comorbidities, medical comorbidities, previous suicidal ideation, previous suicidal behavior, previous self-harm, duration of the current episode, time since initial diagnosis, and number of previous depressive episodes | V0 only |
| Recording and audio traceability variables | Whether the voice recording was obtained, pseudonymized audio file identifier, presence of recording incidents, and type of incident (patient need, clinical need, technical problem, others) | V0–V4 in patients; V0 in controls |
| Clinician-rated and self-reported symptom variables | Depressive symptom severity assessed with MADRS, global clinical impression assessed with ICG, and self-reported depressive symptom burden assessed with PHQ-9 | V0–V4 in patients; V0 in controls |
| Pharmacological treatment variables | Pharmacological treatment received, including total daily dose in mg/24h, dosing interval, administration time, and duration of treatment at the recorded dose | V0–V4 in patients |
| Non-pharmacological treatment and psychotherapy variables | Sleep hygiene measures, diet pattern, regular physical activity, discontinuation of toxic habits, psychotherapy attendance, psychotherapy type, and psychotherapy frequency | V0–V4 in patients |
| Neurostimulation variables | Receipt of electroconvulsive therapy in the current episode, type of electroconvulsive therapy, and other neurostimulation treatments | V0–V4 in patients |

**Table 3.** Multimodal speech- and discourse-derived variables predefined in the VOICE-DEP protocol.

| Domain | Variable / group of variables | Operationalization / measurement |
| --- | --- | --- |
| Acoustic | Pitch | F0 mean, standard deviation and range (Hz). |
|  | Energy / intensity | Mean and standard deviation (dB). |
|  | MFCCs | Coefficients 1–13 summarized by mean and standard deviation. |
|  | Harmonic-to-noise ratio | dB. |
|  | Jitter | Percentage (%). |
|  | Shimmer | Percentage (%). |
|  | Spectral descriptors | Spectral centroid, roll-off and slope (Hz or normalized units). |
|  | Formants | F1–F3 mean frequency and bandwidth (Hz). |
|  | Spectral / prosodic entropy | Dimensionless entropy index. |
|  | Embedding-based acoustic features | High-dimensional vector representations derived from pretrained models (e.g., Wav2Vec2, HuBERT). |
| Paralinguistic | Speech rate / articulation rate / pause duration | Words per second and pause duration in seconds. |
|  | Prosodic range | F0 and intensity variability (Hz, dB, or coefficient of variation). |
|  | Pause ratio | Silence-to-speech proportion (0–1 or %). |
| Linguistic | Sentiment polarity | Continuous score from –1 to +1. |
|  | Negative emotional word ratio | Percentage (%) of negatively valenced words. |
|  | First-person singular pronouns | Percentage (%) or frequency per 100 words. |
|  | Lexical diversity | MTLD, Honor'e and type-token ratio. |
|  | Word count / mean sentence length | Number of words and words per sentence. |
|  | Lexical sentiment variance | Standard deviation of sentiment scores. |
|  | Lexical coherence / cohesion | Cosine similarity based on semantic embeddings. |

### 2.7. Statistical and analytical plan

The analytical strategy of the VOICE-DEP study is designed to support the primary and secondary objectives of the protocol while ensuring methodological transparency, reproducibility, and robustness. In accordance with the study design, the analysis will address three complementary dimensions: (i) descriptive characterization of the study sample and extracted variables, (ii) exploratory cross-sectional association and discrimination analyses between patients with depressive disorder and healthy controls, and (iii) longitudinal intra-individual modeling of changes in speech- and discourse-derived variables in relation to clinical evolution.

First, a descriptive analysis will be performed for all study variables recorded in the case report form. Quantitative variables, including acoustic, paralinguistic, and linguistic features, will be summarized using means and standard deviations or medians and interquartile ranges, as appropriate according to their distribution. Qualitative variables will be described using absolute frequencies and percentages. This descriptive phase will include both baseline variables collected at V0 and longitudinal variables collected across repeated visits in patients.

Second, the association between continuous speech- and discourse-derived biomarkers and clinical severity measures will be explored using Pearson or Spearman correlation coefficients, depending on distributional assumptions. These analyses will focus on the relationship between multimodal voice-derived variables and clinician-rated depressive symptom severity measured with the MADRS, global clinical status measured with the GCI, and self-reported depressive symptom burden measured with the Patient Health Questionnaire-9 (PHQ-9).

For diagnostic discrimination between patients and healthy controls, classification models will be developed and evaluated. The area under the receiver operating characteristic curve (AUC) will be used as the principal performance metric, and additional measures such as odds ratios and their corresponding 95% confidence intervals will be reported where applicable. These analyses will be interpreted as exploratory because the study is a pilot methodological validation study rather than a definitive diagnostic accuracy trial.

Third, a systematic comparison between unimodal and multimodal approaches will be conducted. Unimodal models will include models based exclusively on audio-derived features or text-derived features, whereas multimodal models will combine acoustic, paralinguistic, and linguistic information. This comparison is intended to quantify the added value of multimodal integration beyond individual feature domains and is directly aligned with one of the predefined secondary objectives of the study.

Longitudinal analyses will be used to evaluate the evolution of speech- and discourse-derived biomarkers over time and their association with changes in depressive symptom severity and global clinical status. Because repeated assessments are obtained in the patient group at V0, V1, V2, V3, and V4, these analyses will account for within-subject correlation. Mixed-effects models and, where appropriate, finite-difference or change-score approaches will be used to examine intra-individual trajectories and their relationship with variation in MADRS, ICG, and PHQ-9 scores, as well as with relevant treatment-related variables.

All predictive modeling procedures will use participant-level data splitting in order to preserve the independence of training and test data. Specifically, all recordings from the same participant will be assigned exclusively to either the training or the test partition. This strategy is essential to avoid optimistic bias due to repeated observations from the same individual. In this context, Leave-One-Group-Out cross-validation will be used as an appropriate validation framework for grouped repeated-measures data.

To improve clinical interpretability, feature attribution analyses will be performed using Shapley-based methods, including SHAP values where applicable. These analyses will help identify which acoustic, paralinguistic, and linguistic signals contribute most strongly to model predictions and will support the interpretation of the relative importance of different multimodal domains. No imputation for missing data will be applied. To maximize the data collection, quality control reviews will be conducted after every visit. A flexible time-window for visits and periodic reminder phone calls will be planned to reduce the risk of follow-up losses.

All analyses will be performed using Python v3.x or equivalent validated statistical software. Statistical significance will be considered at a two-sided threshold of p < 0.05.

### 2.8. Safety considerations

The VOICE-DEP study is observational and non-invasive, and does not modify clinical decision-making, treatment prescription, or usual psychiatric follow-up. The only study-specific procedure consists of audio recording a predefined section of the medical interview.

Participants may pause or stop the recording at any time, without providing a reason and without any negative consequence for their medical care. If fatigue, emotional distress, discomfort, or any other difficulty occurs during the interview, the recording may be interrupted or discontinued according to the participant’s preference and the investigator’s clinical judgement.

All assessments will be conducted by qualified clinical staff in a hospital-based psychiatric setting. If clinically relevant worsening, active suicidal ideation, suicidal behavior, severe anxiety, psychotic symptoms, or any other urgent clinical concern is detected, clinical care will take priority over study procedures and the situation will be managed according to usual clinical practice.

Any relevant interruption or incident during the recording, including patient-related, clinical, or technical causes, will be documented in the pseudonymized case report form and considered in data quality assessment.

### 2.9 Ethical considerations and data protection

This protocol has been reviewed and approved by the local Research Ethics Committee, which complies with the international standards of GCP CPMP/ICH/135/95 (Comunidad Foral de Navarra Research Ethics Committee; reference code: 2026.110). The study will be conducted in accordance with the Declaration of Helsinki and with applicable Spanish and European data protection regulations. The approved protocol version is v2, dated 2026/04/27. Recruitment is expected to start in May 2026 and to continue until December 2027. Any substantial amendment to the protocol will be documented, justified and submitted for approval to the Research Ethics Committee before implementation, and will be transparently reported in subsequent publications.

The study is observational and does not modify medical prescription habits or clinical decision-making. It includes review of clinical variables collected in routine practice, administration of the clinical scales specified in the protocol and audio recording of a specific part of the medical interview. No pharmacological or interventional procedure is introduced by the study.

All participants will receive verbal and written information about the study and will provide written informed consent before any study-related procedure. Participants will be informed that participation is voluntary and that they may withdraw at any time without any negative consequence for their subsequent medical care. The informed consent form will include authorization for access to clinical records when required for study objectives.

Participants will be identified using sequential study codes. Case report forms, reports and study communications will be identified using these codes. Only the sponsor, the investigator and authorized members of the research team, the Research Ethics Committee and competent health authorities, where applicable, will have access to identifiable study material.

Pseudonymized voice recordings will be transferred without distortion to secure NNBi servers. Recordings will be stored until 31 December 2029 and destroyed no later than 1 January 2030. NNBi servers are certified according to the Spanish National Security Framework at High level and ISO 27001. Technical and organizational safeguards will include encryption at rest and in transit, role-based access control, the principle of least privilege, robust authentication, environment segregation, access logging, backup encryption, recovery procedures and secure deletion at the end of the retention period.

Results will be reported in aggregate form. Individual-level voice recordings and transcriptions will not be publicly released because of privacy and re-identification risks.

### 2.10 Status and timeline of the study

This study was approved on May 8^th^, 2026. Recruitment and data collection are ongoing and expected to continue until December 2027. Analysis and interpretation of the data are expected for February 2028. Final results report is expected for September 2028.

## 3. Discussion

The present manuscript describes a protocol for a longitudinal, hospital-based study aimed at developing and validating speech-derived digital biomarkers for the detection and monitoring of major depressive disorder. As a protocol paper, this discussion focuses on methodological considerations, anticipated challenges in study implementation, limitations inherent to the design, and plans for dissemination and protocol management, rather than on empirical findings.

### 3.1 Limitations and methodological considerations

Several limitations and anticipated challenges of the proposed protocol should be acknowledged, particularly those inherent to voice-based clinical studies that seek to balance standardization, ecological validity, and translational feasibility.

First, although speech recordings are obtained under controlled hospital conditions, substantial inter-individual variability in speaking style, emotional expressiveness, and engagement during clinical interviews is expected. To balance standardization with clinical realism, a semi-structured, clinician-administered interview protocol is used to elicit naturalistic speech. This improves ecological validity and clinical relevance, but may introduce variability in speech content, duration, and topic coverage, which can in turn affect both acoustic and linguistic measurements.

Second, longitudinal data collection poses practical challenges. Repeated assessments increase participant burden and may lead to missed visits or incomplete follow-up data. To mitigate this risk, follow-up visits are scheduled within flexible time windows and all procedures are designed to be brief and minimally invasive. Nevertheless, some degree of attrition is anticipated and will be explicitly addressed in the analytical strategy.

Third, the planned sample size remains moderate. While the longitudinal structure provides valuable information on intra-individual vocal trajectories, a moderate sample may limit statistical power, increase uncertainty in performance estimates, constrain the exploration of complex interaction effects, and raise the risk of overfitting, particularly for higher-dimensional or multimodal models.

Fourth, technical variability in recording conditions such as background noise or microphone positioning, may influence acoustic measurements. The protocol addresses this by using a standardized recording device and placement, while intentionally avoiding laboratory-grade equipment to maximize clinical applicability. This trade-off prioritizes real-world deployability over maximal signal fidelity and may reduce the signal-to-noise ratio for certain acoustic biomarkers.

Fifth, the study is conducted in a single clinical center and focuses on Spanish-speaking participants, which may restrict external validity across healthcare systems, dialects, or linguistic contexts. However, this focus also addresses a relevant gap in the literature, which has been predominantly centered on English-language datasets.

Finally, clinician-administered scales are used as the primary reference standards, but they are themselves subject to measurement variability and do not constitute an objective ground truth. The present protocol does not aim to replace clinical judgment but rather to complement it by identifying speech-derived markers that correlate with established measures of depressive severity

### 3.2 Dissemination and data sharing

The results derived from this study are intended to be disseminated through peer-reviewed scientific publications and presentations at national and/or international conferences focused on psychiatry, digital health, and biomedical signal processing. Findings will be reported in accordance with relevant reporting guidelines, and emphasis will be placed on transparent description of methods and analytical pipelines.

Where feasible and in compliance with ethical and legal constraints, aggregated results and methodological details will be shared to facilitate replication and comparison with future studies. Individual-level data, including voice recordings and transcriptions, will not be publicly released due to privacy considerations; however, derived features or synthetic representations may be shared upon reasonable request and subject to ethical approval.

### 3.3 Protocol amendments and study termination

Any amendments to the study protocol will be documented, justified, and submitted for approval to the corresponding Research Ethics Committee prior to implementation. Minor procedural adjustments that do not affect participant safety or data integrity will be recorded and reported transparently in subsequent publications.

## Data Availability

No datasets were generated or analysed during the current study. Where feasible and in compliance with ethical and legal constraints, aggregated results and methodological details will be shared to facilitate replication and comparison with future studies. Individual-level data, including voice recordings and transcriptions, will not be publicly released due to privacy considerations however, derived features or synthetic representations may be shared upon reasonable request and subject to ethical approval.

## Supporting information

S1 File: Original protocol.

S2 File: English translation of the original protocol.

S3 File: STROBE checklist of cohort studies (applicable items for a study protocol).

